# Telehealth Equity and Access Communication Skills Pilot Simulation for Practicing Clinicians

**DOI:** 10.1101/2024.04.16.24305892

**Authors:** Christopher J. Nash, Susan E. Farrell, Jossie A. Carreras Tartak, Alexei Wagner, Lea C. Brandt, Emily M. Hayden

**Affiliations:** Department of Emergency Medicine, Duke University Hospital, Durham, North Carolina, USA; Harvard Medical School, Boston, Massachusetts, USA; Department of Emergency Medicine, Beth Israel Deaconess Medical Center, Boston, Massachusetts, USA; Department of Emergency Medicine, Brigham and Women’s Hospital, Boston, Massachusetts, USA; Center for Health Ethics, University of Missouri School of Medicine, Columbia, Missouri, USA; Department of Emergency Medicine, Massachusetts General Hospital, Boston, Massachusetts, USA

## Abstract

**Objectives:** This pilot study evaluated a telehealth training simulation program for practicing clinicians, specifically focused on addressing patient issues of equity and access to healthcare via improving telehealth communication.

**Methods:** Participants participated in a one-hour simulation experience with two cases. Performance was assessed pre- and post-intervention using a checklist measuring communication domains related to equity and access in telehealth. Participant satisfaction was secondarily measured via survey.

**Results:** Results showed measurable gains in clinicians’ abilities to effectively incorporate equity and access communication skills. Participants found the session useful and recommended the training experience.

**Conclusions:** The findings of this pilot study highlight the potential of simulation-based telehealth training for practicing clinicians, emphasizing clinicians’ attention to patients’ equitable access to healthcare. Future studies should aim to explore the durability of learning and investigate the generalizability of this training approach to other telehealth competencies and settings.

## Introduction

As telehealth becomes more integrated into healthcare delivery [1–4], it is increasingly important that healthcare professionals learn the necessary skills to effectively care for patients in the virtual environment. Healthcare delivery via telehealth presents distinct challenges from the traditional care environment [5]. The Association of American Medical Colleges (AAMC) created core competencies defining specific skills including conducting a virtual physical examination, troubleshooting technological failures, and communicating through a virtual connection [6–8]. The AAMC telehealth competencies serve as the best guide to-date for designing and implementing curricula for telehealth training and competency evaluation for medical providers.

Despite the growing need to train medical professionals in virtual care, as of 2021-22, less than sixty percent of medical schools and nursing schools included any formal training in virtual healthcare delivery [9,10], and even fewer physician assistant (PA) programs incorporated formal telehealth curricula [11]. Unfortunately, those programs that include telehealth training implement their courses as electives and without robust evaluation of educational outcomes [12]. This lack of undergraduate preparation for telehealth care means that most telehealth practitioners have never been formally taught how to best care for their patients remotely [13,14], creating a training gap that should be addressed urgently.

Simulation-based training has proven to be an effective educational method in healthcare, as clinicians can practice and refine their communication skills in a risk-free environment, and standardized patient (SP) actors have been increasingly utilized to conduct realistic simulated telehealth encounters for training and assessment [15–21]. The use of standardized patients to deliver feedback as a primary component of education has been extensively studied and is known to be an effective technique for adult learning in simulated environments, including in telehealth simulation [22–25]. However, most descriptions of telehealth training in the literature that utilize SP actors involve undergraduate trainees—depictions of SP-based training for the practicing clinician are growing but are currently scant [22,26–28], and there is a lack of specific, evidence-based information in the literature about communication in telehealth care for practicing clinicians [29]. Our team’s previous experience with video-based simulated telehealth encounters demonstrated that practicing clinicians believe that this type of program builds confidence and skills in the use of the telehealth modality [22]. We focused on communication skills for this study because we recognized that there is significant risk of perpetuating inequity in healthcare, and telehealth may add an additional layer of complexity for these interactions that prior formal communications training may not have adequately addressed [30–34]. Furthermore, to date no studies have focused on training programs specifically oriented to incorporate issues of equity and access in the telehealth—a key domain within the AAMC telehealth competencies that is important yet currently underexplored [6]. The incorporation of telehealth in clinical practice has the potential to exacerbate inequity; our team believes that formal, intentional educational efforts to combat this are needed [30,32].

This pilot study sought to address the telehealth communications training gap by creating a simulation experience for practicing clinicians, suitable for both physicians and advanced practice practitioners (APPs). We aimed to assess the efficacy of an SP-delivered educational experience in which the SP provided both the portrayal of the patient and the generation and delivery of feedback to the practicing clinicians as an educational intervention. Our primary outcome was the practicing clinicians’ checklist-based performance on AAMC-aligned telehealth competencies, specifically centered on Domains II and III (equity, access, and communication) [8]. Secondarily, we captured the participants’ perceptions of knowledge acquisition via pre- and post-session survey.

## Methods

### Study Design

This study was a prospective interventional study that received pilot funding from the AAMC Telehealth Equity Catalyst (TEC) grant. Utilizing Kolb’s Experiential Learning conceptual framework [35–37], an SP-led one-hour simulation-based learning experience was designed consisting of two cases. Learning objectives (S1 Appendix) for the sessions were designed to map to the AAMC telehealth competencies [8]. Each participant completed both cases (A and B), in a cross-over design with half doing Case A first and half doing Case B first. Standardized patient feedback was given to participants after each case by the SP. Participants were surveyed before and after the experience, and their performance was recorded in performance checklists.

### Setting and Participants

Eligible participants were physicians or APPs within a fourteen-hospital medical system serving a large urban area in the Northeastern United States practicing in generalist specialties, including emergency medicine, virtual urgent care, internal medicine, and primary care. We did not include pediatrics in this study for consistency of the simulated telehealth patient encounters. Subjects participated on a voluntary basis and were scheduled at times convenient to their schedules. There were no exclusion criteria. Study participants were recruited via email between January and April 2023. The recruitment email outlined the study, encompassing both the potential risks and benefits. Additionally, it reminded participants of their right to withdraw from the study at any point. Data was collected contemporaneously. We aimed for the recruitment of 30 individuals for participation to comply with budgetary limitations in this study. Participants’ consent was implied by participation in the study, and the requirement for formal written documentation of consent was waived by the institutional review board (IRB). This study was approved by the IRB at Massachusetts General Hospital (Agreement number 2022A006072).

### Case Design

We designed two cases (S2 Appendix and S3 Appendix) suitable to a generalist / urgent care environment, intentionally crafted by our study team to surface issues of communication and equity in healthcare access. Case A was a case of a 56-year-old person with hypertension seeking care for headaches, but with additional life stressors including juggling the potential loss of a job, caring for her grandchildren, and participating in online school. Case B was a 56-year-old person with a history of asthma who had been missing work due to frequent exacerbations, but with difficulty affording medication refills. The cases were written and iterated by experts in telehealth, standardized patient case design, ethics, and diversity, equity, and inclusion.

Prior to the first live session with participants, we held a training session with the SP actors to ensure consistency in the actor portrayal. Additionally, as the SPs were expected to complete checklists and provide feedback after each simulated encounter, this activity was modeled and practiced by the SPs. We reviewed the cases with the SPs for clarity and adjusted their timing to fit in the ten minutes allotted for each simulation.

### Data Sources and Instrument Design

Prior to the simulation sessions, participants received learning objectives and a pre-session survey (S4 Appendix). Surveys were administered using REDCap (Vanderbilt University, Nashville, TN). There were no other pre-session requirements for the session, including no pre-session didactics, minimizing the time requirements for the clinicians who volunteered to participate in this study outside of regular work hours. Sessions took place via Microsoft Teams (Microsoft Corporation, Redmond, WA).

Each session included two participants and two trained SPs. Each encounter lasted approximately 10 minutes, immediately followed by 7 minutes during which the SP delivered immediate feedback to the participant. We created a learning objective-aligned checklist to assess performance and communication skills of the participants that was modified from the Kalamazoo Essential Elements Communications Checklists [38,39]. This checklist was chosen as a template for our project because it has previously been validated, including in modified forms [40–42]. The checklist contained a total of 22 assessment items (S5 Appendix). Participants’ performance was assessed in each of their two cases.

The SP completed the checklist in REDCap (Vanderbilt University, Nashville, TN) simultaneous to or immediately after providing feedback at the end of each case. This allowed feedback to be aligned with learning objectives and, therefore, the AAMC competencies [8]. Participants then performed their second 10-minute case, followed again by feedback. After the session, participants completed a post-session survey (S6 Appendix). The surveys were designed to gauge participants’ self-perception of learning objective-aligned skills as a measure of growth, as well as demographic data. A crossover design was utilized to reduce the risk that measured improvement could be due to unmeasured differences in difficulty between the two cases.

### Data Analysis

Data was analyzed using Stata (StataCorp, College Station, Texas, version 17) and Microsoft Excel (Microsoft Corporation, Redmond, WA). Descriptive statistics were generated to understand the demographics of the study participants. Paired T-tests were used to test for differences in pre- and post-session survey responses and to evaluate differences in participants’ checklist performance between cases one and two.

## Results

A total of 30 clinicians participated in the study (Table 1). Participants included six physicians (20%) and 24 advanced practice providers (NPs or PAs). Participants ranged in experience (0-20+ years out of training), and approximately half had participated in telehealth in the past (15, one missing response). All participants (100%) completed all pre and post surveys, and all performance checklists (100%) were completed by the SPs.

**Table 1:**
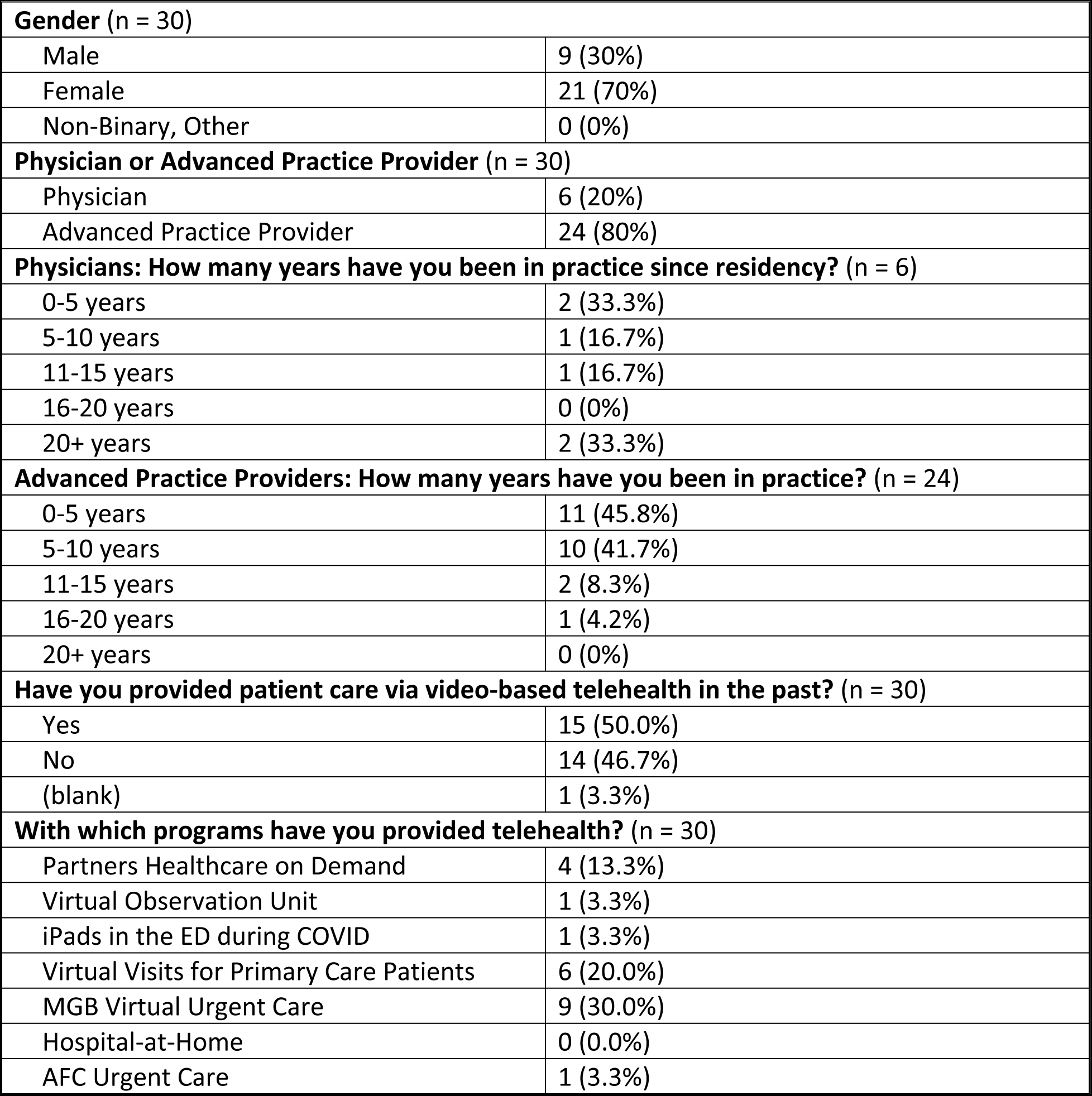
Participant Characteristics.

### Primary Objectives

Our primary objective was performance changes between cases as measured by the checklist. Overall, baseline performance for most checklist items for their first case was quite high, and in many cases all 30 participants met the correctly performed checklist items in both of their cases (Table 2). Some item ratings demonstrated statistically significant improvements after receiving SP feedback on the first case. Specifically, improvement in performance was seen for “ensures my privacy by making sure that my space is private for me” (p = 0.0226), “ensures my privacy by making sure and indicating they are in a private space for their conversation (e.g., nobody else can hear our conversation on their end)” (p < 0.01), and “ensures that I have access to resources that will support my post-encounter care” (p = 0.0117).

**Table 2:**
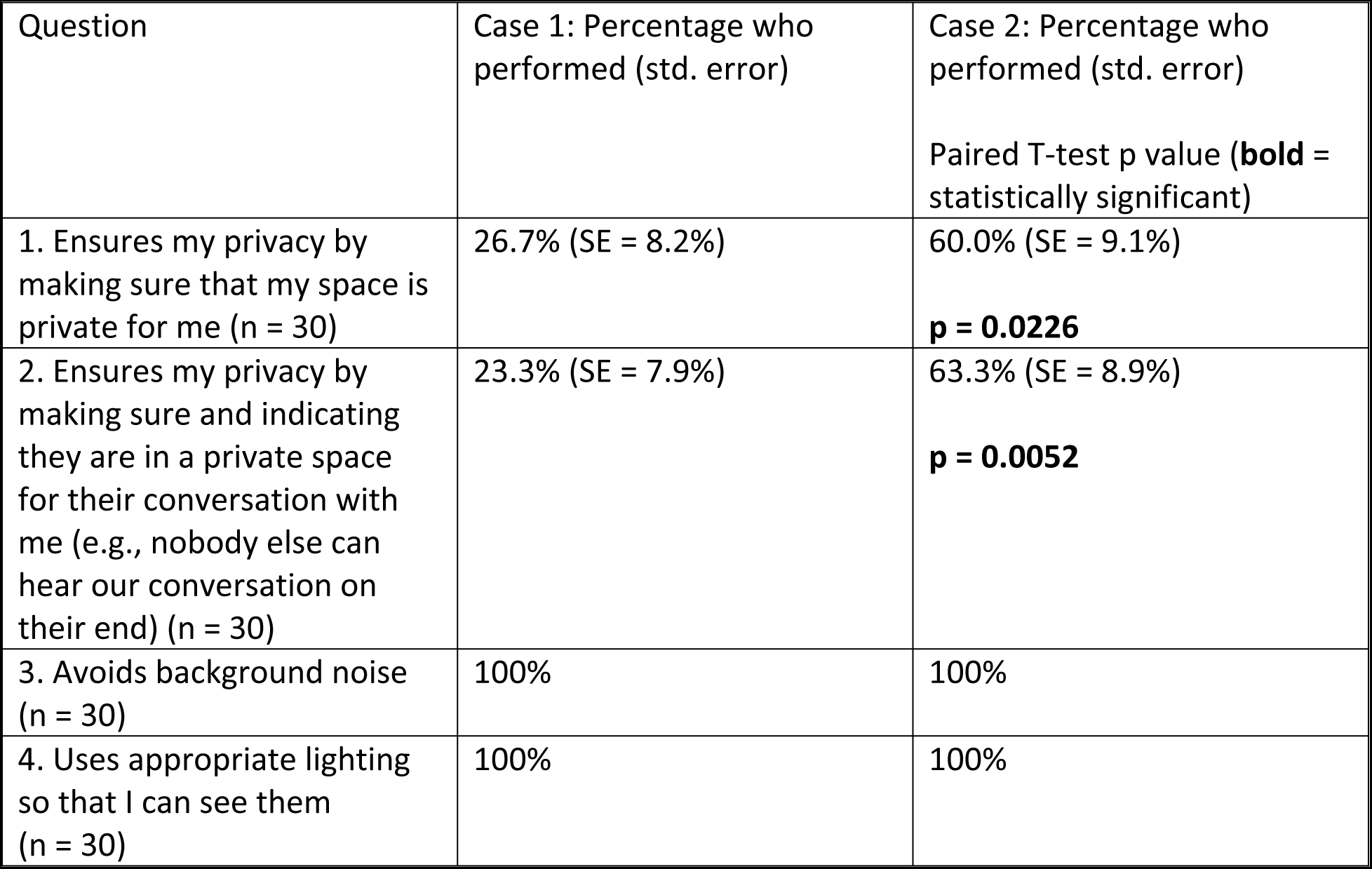
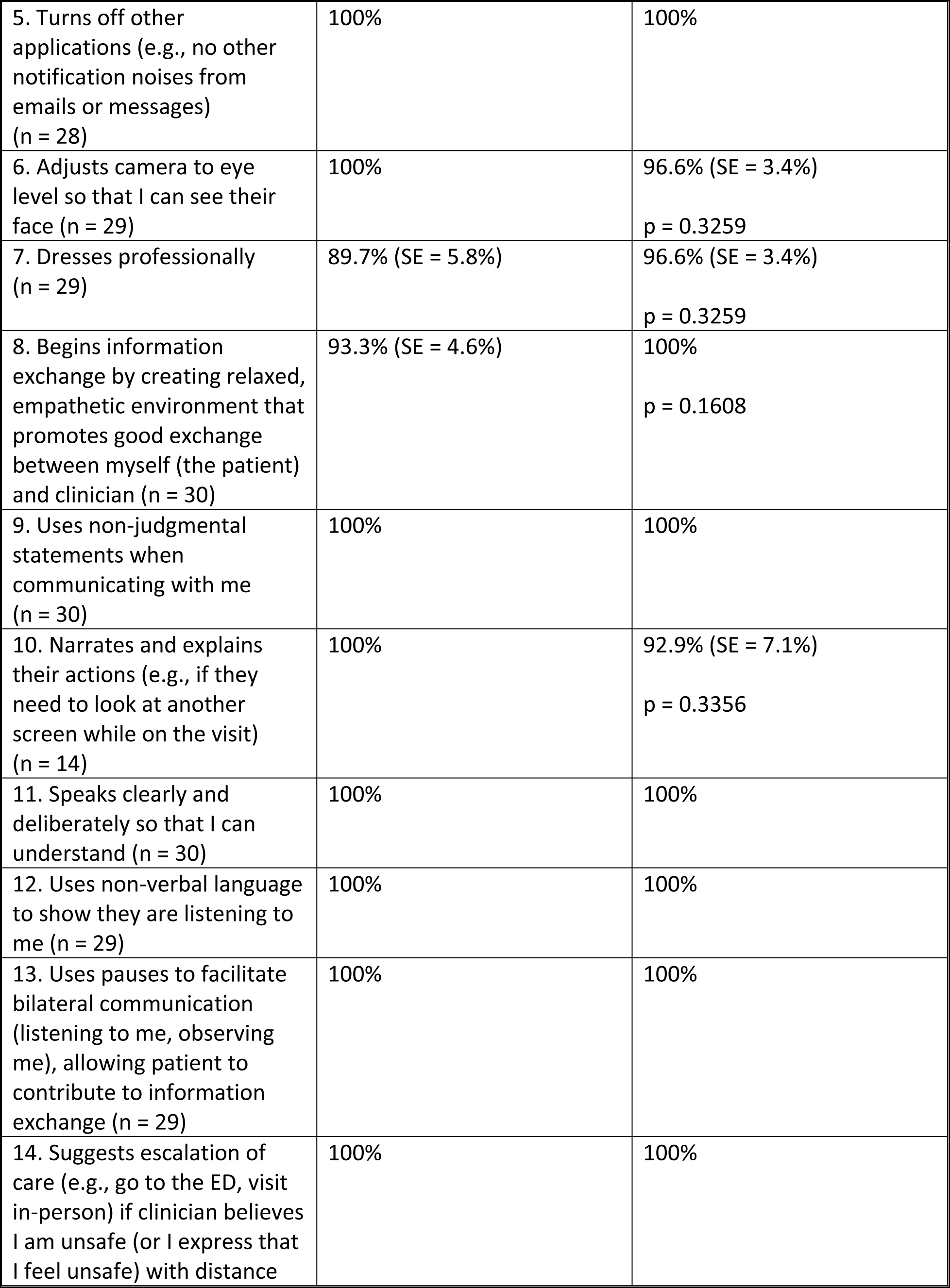
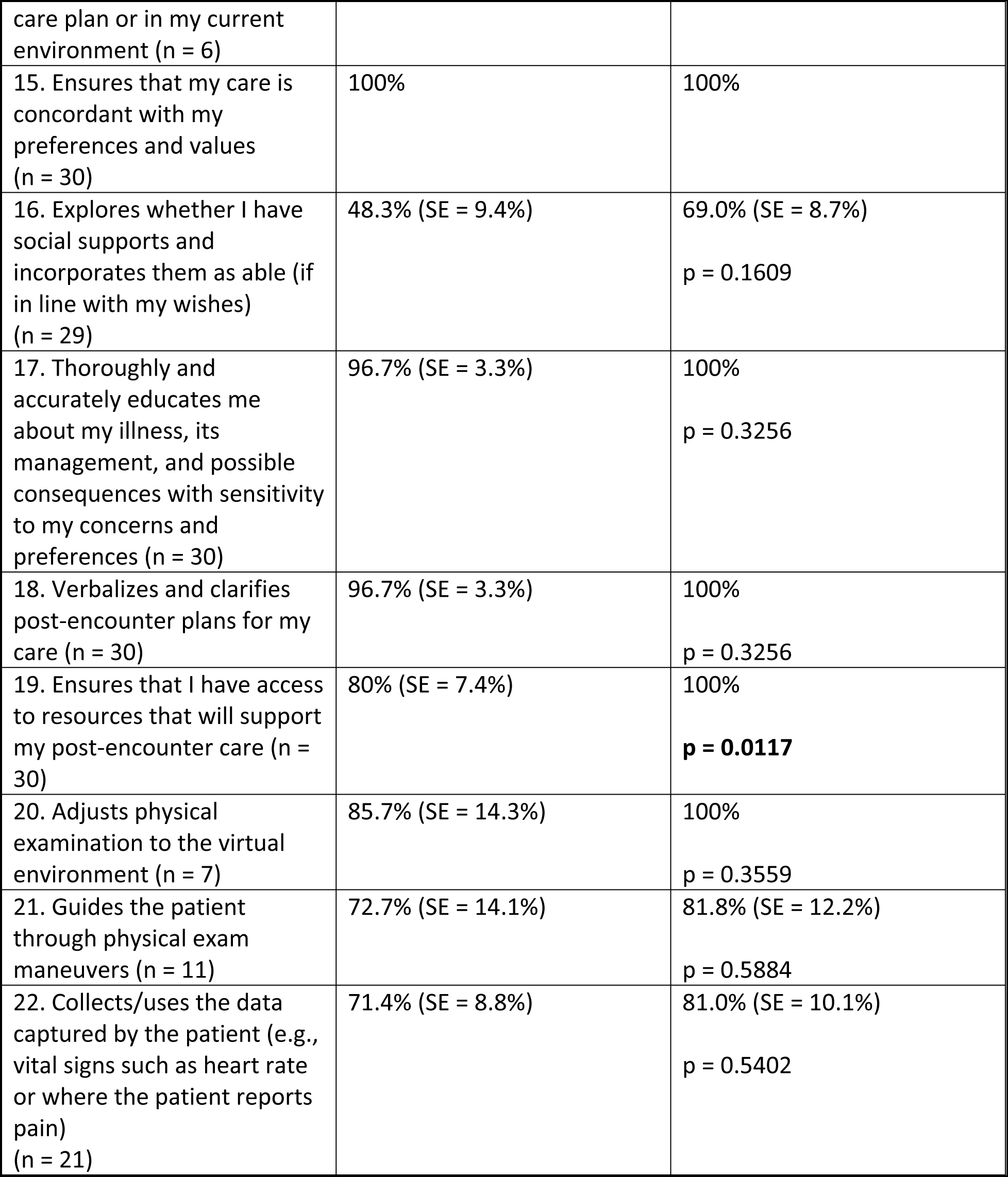
Checklist Performance by Case.

### Secondary Objectives

Overall, participants reported that they would recommend this training experience, with a mean of 8.8 (SD 1.49) on a Likert scale from 0 (Not at all likely) to 10 (Extremely likely). Participants’ responses to every survey question demonstrated a statistically significant improvement in self-perceived performance/skill level in all learning-objective aligned items (Table 3).

**Table 3:**
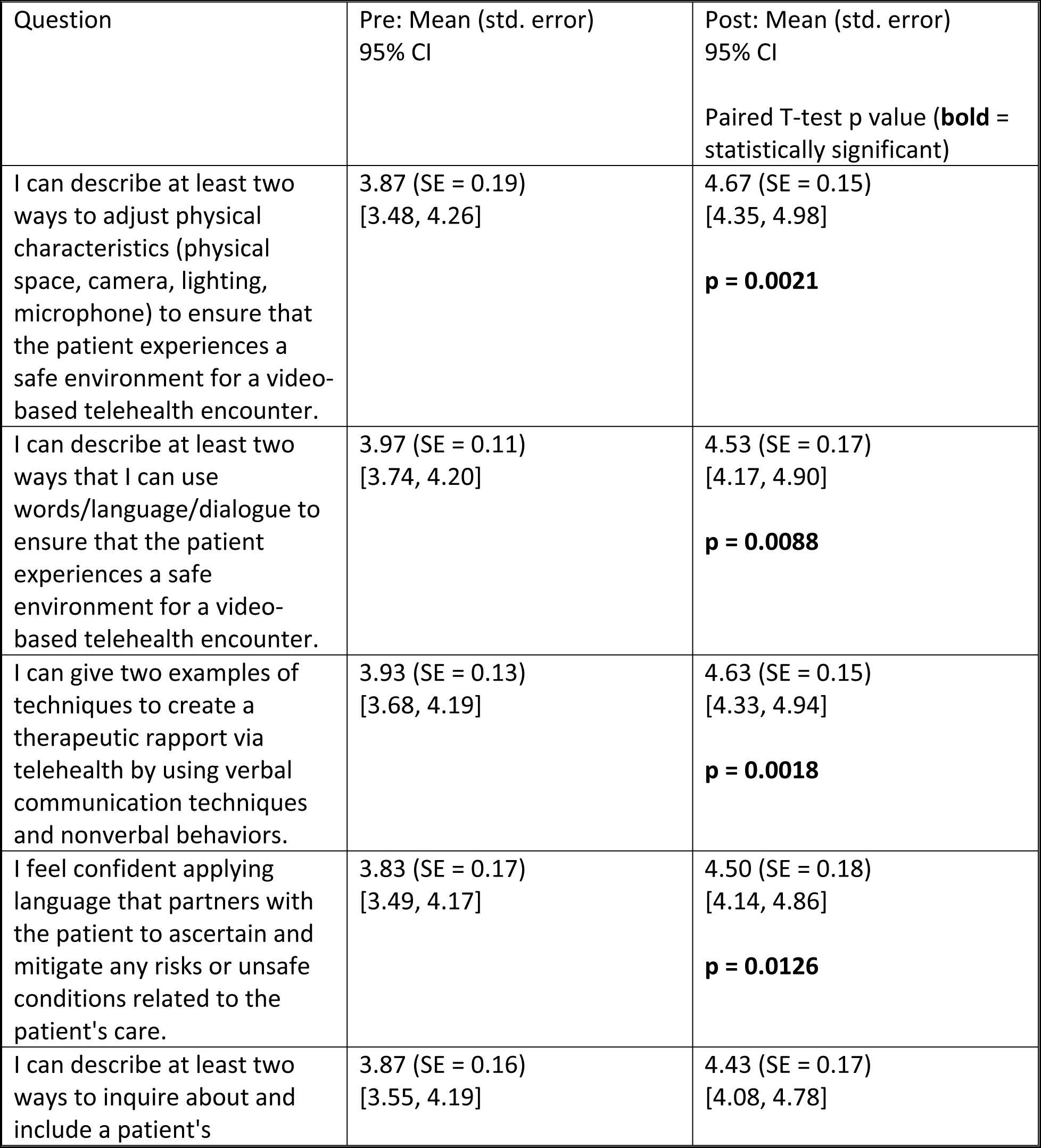
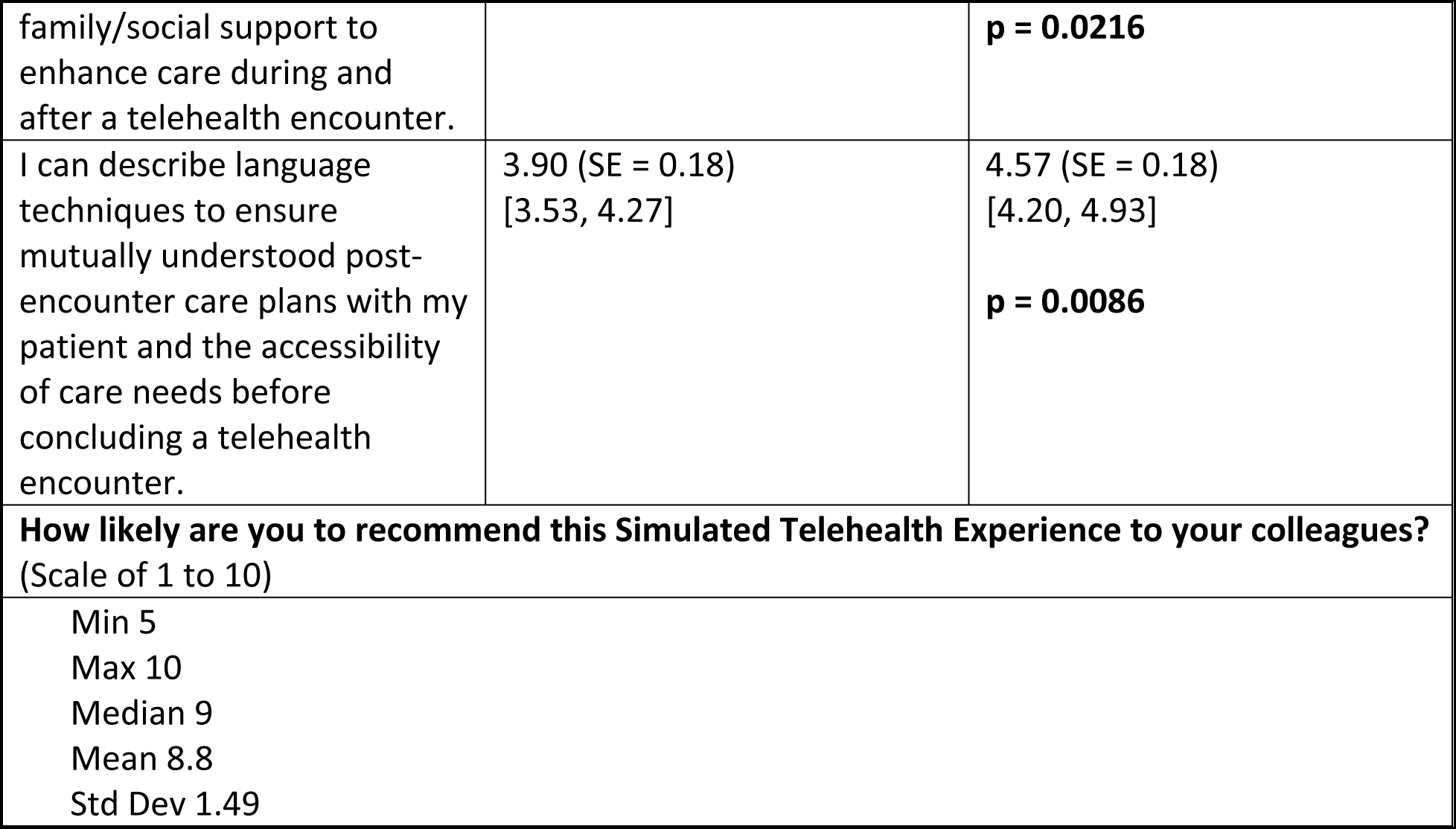
Participants’ Self-Perceived Performance Pre- and Post-Session Attitudes.

## Discussion

As telehealth education continues to assume a more central role in health professions education, studies that demonstrate evidence for effective training are timely and needed. In this pilot study, we conducted a training program for 30 practicing clinicians. We found that the use of SP-generated feedback as a primary educational strategy resulted in improved performance as measured on learning-objective aligned checklists, with improvement in multiple domains related to equity and access in healthcare delivery reaching statistical significance. The participants rated the learning experience highly and, when surveyed, endorsed an improvement in confidence and skills for all measured learning objectives. We believe this to be the first study to demonstrate improved telehealth communication performance via telehealth simulated encounters for patient equity and access in healthcare delivery. This is an important area for future research that should be replicated and expanded upon to raise telehealth as a model for providing healthcare to patient populations who may otherwise have limited access. It is possible that this finding could be generalized from practicing clinicians to those in training.

One key aspect we sought to assess in our study was whether a simulation session without pre-session didactics could be effective. As a pilot, it was deemed beyond the scope of the study to develop a series of didactics, but it was also of interest to our team to assess if a shorter training approach could engender measurable learning and change for the learners. Traditionally, pre-session didactics have been included as a part of a module or course [43–45]; however, these studies typically are intended for medical trainees and not practicing clinicians [15,16,18,20,43–45]. Our study adds to the literature base for telehealth training for practicing clinicians. Typically courses with pre-session didactics require more time and may be less feasible for practicing clinicians who have limited training time. Our study suggests that, at least in some circumstances or for some competencies such as those focused on equity and access, simulation sessions that include focused SP feedback can stimulate behavior change with less time burden. Future research could continue to investigate short simulation sessions using a variety of case complexities as a mechanism of instruction for busy practicing clinicians.

This study demonstrates the feasibility of an actor-run session for telehealth communication training. Any software that allows for video calls, such as Microsoft Teams (Microsoft Corporation, Redmond, WA) or Zoom (Zoom Video Communications, San Jose, CA) could be utilized for sessions at low or no cost. The primary resource for carrying out this type of session is the availability of funds to pay SPs, a cost that should be similar to other simulation sessions that utilize SP services. In our study, the trained SPs proved able to deliver feedback that improved performance on the participants’ subsequent case. Thus, it is possible that trained SPs may be capable of teaching this skillset with reduced faculty or instructor involvement, enhancing the efficiency of resource use. The use of SP feedback as a modality for instruction has been well described in the literature [22,23,23–25,46], but this is the first study to our knowledge that demonstrates improved telehealth performance via this approach. In addition, it appears to be the first to focus on training in patient equity and access topics in telehealth. Future studies may further investigate or validate this finding, and to determine under which circumstances SPs alone may be able to teach telehealth communication skills.

Given the scarcity of educational literature evaluating teaching methods for telehealth communication for practicing clinicians, innovative and evidence-based techniques are urgently necessary. Our study shows that communication skills for patient equity and access in healthcare can be taught efficiently to health professionals. This method of training may be generalizable to other telehealth competencies, such as technology failures or legal / ethical issues in telehealth, suggesting that future research should be directed to this aim.

### Limitations

As a pilot study, the sample size was small and used no control group, limiting generalizability to other settings. Since participation in this study was voluntary, clinicians who were particularly interested or who felt particularly inexperienced in telehealth may have volunteered, contributing to selection bias, which may have increased the measured effect size of the intervention. Although we sought representation from multiple generalist specialties, these results may not be generalizable to all specialties or non-academic urban medical centers. Limited resources prevented a measurement of the durability of learning, an area to direct future study. Additionally, the study did not include a formal needs assessment, which could have provided further insights into the specific educational needs of practicing clinicians in telehealth communication. Finally, although we feel that the use of a modified Kalamazoo Essential Elements Communications Checklists was logical and similar approaches have been taken in the past [18], any modification of an instrument threatens its validity.

## Conclusion

This pilot study underscores the potential of simulation-based telehealth training for practicing clinicians. The findings suggest that such training can effectively enhance telehealth communication skills and address issues of equity and access in virtual healthcare delivery. The study also demonstrates participant satisfaction with actor-run sessions, which could be a cost-effective and time-efficient approach to telehealth training. However, the study’s limitations, including its small sample size and lack of a control group, highlight the need for further research. Future studies should aim to validate these findings, explore the durability of learning, and investigate the generalizability of this training approach to other telehealth competencies and settings.

## Data Availability

All relevant data are within the manuscript and its Supporting Information files.

## Acknowledgements

The authors thank the Association of American Medical Colleges for their help and guidance in conducting this research.

## Supporting Information

**S1 Appendix.** Learning Objectives

**S2 Appendix:** Case A

**S3 Appendix:** Case B

**S4 Appendix:** Pre-Session Survey

**S5 Appendix:** Standardized Patient Checklist

**S6 Appendix:** Post-Session Survey

## Notes

### Competing Interest Statement

The authors have declared no competing interest.

### Funding Statement

Yes

### Author Declarations

This study was approved by the institutional review board at Massachusetts General Hospital (Agreement number 2022A006072).

## References

1. Hollander JE, Carr BG. Virtually Perfect? Telemedicine for Covid-19. N Engl J Med. 2020;382: 1679–1681. doi:10.1056/NEJMp2003539

2. Bashshur RL, Howell JD, Krupinski EA, Harms KM, Bashshur N, Doarn CR. The Empirical Foundations of Telemedicine Interventions in Primary Care. Telemed J E-Health Off J Am Telemed Assoc. 2016;22: 342–375. doi:10.1089/tmj.2016.0045

3. Koonin LM, Hoots B, Tsang CA, Leroy Z, Farris K, Jolly T, et al. Trends in the Use of Telehealth During the Emergence of the COVID-19 Pandemic - United States, January-March 2020. MMWR Morb Mortal Wkly Rep. 2020;69: 1595–1599. doi:10.15585/mmwr.mm6943a3

4. Strazewski L. Telehealth’s post-pandemic future: Where do we go from here? In: American Medical Association [Internet]. 7 Sep 2020 [cited 21 Sep 2023]. Available: https://www.ama-assn.org/practice-management/digital/telehealth-s-post-pandemic-future-where-do-we-go-here

5. van Galen LS, Wang CJ, Nanayakkara PWB, Paranjape K, Kramer MHH, Car J. Telehealth requires expansion of physicians’ communication competencies training. Med Teach. 2019;41: 714–715. doi:10.1080/0142159X.2018.1481284

6. Galpin K, Sikka N, King SL, Horvath KA, Shipman SA, AAMC Telehealth Advisory Committee. Expert Consensus: Telehealth Skills for Health Care Professionals. Telemed J E-Health Off J Am Telemed Assoc. 2021;27: 820–824. doi:10.1089/tmj.2020.0420

7. Sharma R, Nachum S, Davidson KW, Nochomovitz M. It’s not just FaceTime: core competencies for the Medical Virtualist. Int J Emerg Med. 2019;12: 8. doi:10.1186/s12245-019-0226-y

8. AAMC. Telehealth competencies across the learning continuum. [cited 21 Sep 2023]. Available: https://collections.nlm.nih.gov/catalog/nlm:nlmuid-9918504887606676-pdf

9. Khullar D, Mullangi S, Yu J, Weems K, Shipman SA, Caulfield M, et al. The state of telehealth education at U.S. medical schools. Healthc Amst Neth. 2021;9: 100522. doi:10.1016/j.hjdsi.2021.100522

10. Eckhoff DO, Guido-Sanz F, Anderson M. Telehealth across nursing education: Findings from a national study. J Prof Nurs. 2022;42: 308–314. doi:10.1016/j.profnurs.2022.07.013

11. Fleming S, Gordes KL, Cawley JF, Kulo V, Hagar E, Jun H-J, et al. Advancing Telehealth Competency in Physician Assistant Education: Stakeholder Perspectives and a Curricular Model for PA Programs. J Physician Assist Educ. 2022;33: 353. doi:10.1097/JPA.0000000000000461

12. Car LT, Kyaw BM, Panday RSN, Kleij R van der, Chavannes N, Majeed A, et al. Digital Health Training Programs for Medical Students: Scoping Review. JMIR Med Educ. 2021;7: e28275. doi:10.2196/28275

13. DuBose-Morris R, Coleman C, Ziniel SI, Schinasi DA, McSwain SD. Telehealth Utilization in Response to the COVID-19 Pandemic: Current State of Medical Provider Training. Telemed E-Health. 2022;28: 1178–1185. doi:10.1089/tmj.2021.0381

14. Garber K, Gustin T. Telehealth Education: Impact on Provider Experience and Adoption. Nurse Educ. 2022;47: 75–80. doi:10.1097/NNE.0000000000001103

15. Mulcare M, Naik N, Greenwald P, Schullstrom K, Gogia K, Clark S, et al. Advanced Communication and Examination Skills in Telemedicine: A Structured Simulation-Based Course for Medical Students. MedEdPORTAL. 16: 11047. doi:10.15766/mep_2374-8265.11047

16. Cantone RE, Palmer R, Dodson LG, Biagioli FE. Insomnia Telemedicine OSCE (TeleOSCE): A Simulated Standardized Patient Video-Visit Case for Clerkship Students. MedEdPORTAL. 15: 10867. doi:10.15766/mep_2374-8265.10867

17. Belakovskiy A, Jones EK. Telehealth and Medical Education. Prim Care. 2022;49: 575–583. doi:10.1016/j.pop.2022.04.003

18. Farrell SE, Junkin AR, Hayden EM. Assessing Clinical Skills Via Telehealth Objective Standardized Clinical Examination: Feasibility, Acceptability, Comparability, and Educational Value. Telemed E-Health. 2022;28: 248–257. doi:10.1089/tmj.2021.0094

19. Dahmen L, Linke M, Schneider A, Kühl SJ. Medical students in their first consultation: A comparison between a simulated face-to-face and telehealth consultation to train medical consultation skills. GMS J Med Educ. 2023;40: Doc63. doi:10.3205/zma001645

20. Murphy E, Stein A, Pahwa A, McGuire M, Kumra T. Improvement of Medical Student Performance in Telemedicine Standardized Patient Encounters Following an Educational Intervention. Fam Med. 2023;55: 400–404. doi:10.22454/FamMed.2023.523442

21. Lesselroth B, Monkman H, Liew A, Palmer R, Crosby K, Kelly D, et al. Simulating Telemedicine, Medication Reconciliation, and Social Determinants: A Novel Instructional Approach to Health Systems Competencies. Stud Health Technol Inform. 2024;310: 1201– 1205. doi:10.3233/SHTI231155

22. Hayden EM, Nash CJ, Farrell SE. Simulated video-based telehealth training for emergency physicians. Front Med. 2023;10. Available: https://www.frontiersin.org/articles/10.3389/fmed.2023.1223048

23. Doyle Howley L, Martindale J. The Efficacy of Standardized Patient Feedback in Clinical Teaching: A Mixed Methods Analysis. Med Educ Online. 2004;9: 4356. doi:10.3402/meo.v9i.4356

24. Berenson LD. Standardized Patient Feedback: Making It Work Across Disciplines. J Allied Health. 2012;41.

25. Lin EC-L, Chen S-L, Chao S-Y, Chen Y-C. Using standardized patient with immediate feedback and group discussion to teach interpersonal and communication skills to advanced practice nursing students. Nurse Educ Today. 2013;33: 677–683. doi:10.1016/j.nedt.2012.07.002

26. Wallach A, McCrickard M, Eliasz KL, Hochman K. An experiential faculty orientation to set communication standards. Med Educ. 2019;53: 512–513. doi:10.1111/medu.13867

27. Sartori DJ, Lakdawala V, Levitt HB, Sherwin JA, Testa PA, Zabar SR. Standardizing Quality of Virtual Urgent Care: Using Standardized Patients in a Unique Experiential Onboarding Program. MedEdPORTAL. 18: 11244. doi:10.15766/mep_2374-8265.11244

28. Cruz-Panesso I, Tanoubi I, Drolet P. Telehealth Competencies: Training Physicians for a New Reality? Healthc Basel Switz. 2023;12: 93. doi:10.3390/healthcare12010093

29. Noronha C, Lo MC, Nikiforova T, Jones D, Nandiwada DR, Leung TI, et al. Telehealth Competencies in Medical Education: New Frontiers in Faculty Development and Learner Assessments. J Gen Intern Med. 2022;37: 3168–3173. doi:10.1007/s11606-022-07564-8

30. Samuels-Kalow M, Jaffe T, Zachrison K. Digital disparities: designing telemedicine systems with a health equity aim. Emerg Med J EMJ. 2021;38: 474–476. doi:10.1136/emermed-2020-210896

31. Drossman DA, Chang L, Deutsch JK, Ford AC, Halpert A, Kroenke K, et al. A Review of the Evidence and Recommendations on Communication Skills and the Patient-Provider Relationship: A Rome Foundation Working Team Report. Gastroenterology. 2021;161: 1670–1688.e7. doi:10.1053/j.gastro.2021.07.037

32. Rising KL, Kemp M, Leader AE, Chang AM, Monick AJ, Guth A, et al. A Prioritized Patient-Centered Research Agenda to Reduce Disparities in Telehealth Uptake: Results from a National Consensus Conference. Telemed Rep. 2023;4: 387–395. doi:10.1089/tmr.2023.0051

33. Gustavson AM, Lewinski AA, Fitzsimmons-Craft EE, Coronado GD, Linke SE, O’Malley DM, et al. Strategies to Bridge Equitable Implementation of Telehealth. Interact J Med Res. 2023;12: e40358. doi:10.2196/40358

34. Guizado de Nathan G, Shaw LK, Doolen J. Social Determinants of Health: A Multilingual Standardized Patient Case to Practice Interpreter Use in a Telehealth Visit. MedEdPORTAL J Teach Learn Resour. 2023;19: 11364. doi:10.15766/mep_2374-8265.11364

35. Gaba DM. The future vision of simulation in health care. BMJ Qual Saf. 2004;13: i2–i10. doi:10.1136/qshc.2004.009878

36. Taylor DCM, Hamdy H. Adult learning theories: Implications for learning and teaching in medical education: AMEE Guide No. 83. Med Teach. 2013;35: e1561–e1572. doi:10.3109/0142159X.2013.828153

37. Kolb DA. Experiential Learning: Experience as the Source of Learning and Development. FT Press; 2014.

38. Makoul G. Essential Elements of Communication in Medical Encounters: The Kalamazoo Consensus Statement. Acad Med. 2001;76: 390.

39. Calhoun AW, Rider EA, Meyer EC, Lamiani G, Truog RD. Assessment of communication skills and self-appraisal in the simulated environment: feasibility of multirater feedback with gap analysis. Simul Healthc J Soc Simul Healthc. 2009;4: 22–29. doi:10.1097/SIH.0b013e318184377a

40. Joyce BL, Steenbergh T, Scher E. Use of the kalamazoo essential elements communication checklist (adapted) in an institutional interpersonal and communication skills curriculum. J Grad Med Educ. 2010;2: 165–169. doi:10.4300/JGME-D-10-00024.1

41. Porcerelli JH, Brennan S, Carty J, Ziadni M, Markova T. Resident Ratings of Communication Skills Using the Kalamazoo Adapted Checklist. J Grad Med Educ. 2015;7: 458–461. doi:10.4300/JGME-D-14-00422.1

42. Brown SD, Rider EA, Jamieson K, Meyer EC, Callahan MJ, DeBenedectis CM, et al. Development of a Standardized Kalamazoo Communication Skills Assessment Tool for Radiologists: Validation, Multisource Reliability, and Lessons Learned. Am J Roentgenol. 2017;209: 351–357. doi:10.2214/AJR.16.17439

43. Smith TS, Watts P, Moss JA. Using Simulation to Teach Telehealth Nursing Competencies. J Nurs Educ. 2018;57: 624–627. doi:10.3928/01484834-20180921-10

44. Wong R, Ng P, Spinnato T, Taub E, Kaushal A, Lerman M, et al. Expanding Telehealth Competencies in Primary Care: A Longitudinal Interdisciplinary Simulation to Train Internal Medicine Residents in Complex Patient Care. J Grad Med Educ. 2020;12: 745–752. doi:10.4300/JGME-D-20-00030.1

45. Eckhoff DO, Diaz DA, Anderson M. Using Simulation to Teach Intraprofessional Telehealth Communication. Clin Simul Nurs. 2022;67: 39–48. doi:10.1016/j.ecns.2022.03.006

46. Beaird G, Nye C, Thacker LR. The Use of Video Recording and Standardized Patient Feedback to Improve Communication Performance in Undergraduate Nursing Students. Clin Simul Nurs. 2017;13: 176–185. doi:10.1016/j.ecns.2016.12.005

